# What can trends in hospital deaths from COVID-19 tell us about the progress and peak of the pandemic? pandemic? An analysis of death counts from England announced up to 25 April 2020

**DOI:** 10.1101/2020.04.21.20073049

**Authors:** David A Leon, Christopher I Jarvis, Anne Johnson, Liam Smeeth, Vladimir M Shkolnikov

## Abstract

**Background:** Reporting of daily hospital COVID-19 deaths in the UK are promoted by the government and scientific advisers alike as a key metric for assessing the progress in the control of the epidemic. These data, however, have certain limitations, among which one of the most significant concerns the fact that the daily totals span deaths that have occurred between 1 and 10 days or more in the past.

**Data and methods:** We obtained daily data published published by NHS England up to and including April 25 in the form of Excel spreadsheets in which deaths counts are presented by date of death according to age and region. Simple descriptive analyses were conducted and presented in graphical and tabular form which were aimed at illustrating the biases inherent in focussing on daily counts regardless of when the deaths occurred. We then looked at how a less biased picture could be obtained by looking at trends in death counts stratifying by individual period of delay in days between occurrence of death and when the death was included in the daily announcement.

**Findings:** The number of hospital COVID-19 deaths announced daily overestimates the maximum number of deaths actually occurring so far in the epidemic in the UK, and also obscures the pattern of decline in deaths. Taking account of reporting delays suggests that for England as a whole a peak in hospital COVID-19 deaths may have been reached on April 8 with a subsequent gradual decline suggested. The same peak is also seen among those aged 60-79 and 80+, although there is slightly shallower decline in the oldest age group (80+ years). Among those aged 40-59 years a later peak on April 11 is evident. London shows a peak on April 8 and a clearer and steeper pattern of subsequent decline compared to England as a whole.

**Interpretation:** Analyses of mortality trends must take account of delay, and in communication with the public more emphasis should be placed on looking at trends based on deaths that occurred 5 or more days prior to the announcement day. The slightly weaker decline seen at age 80+ may reflect increased hospitalisation of people from care homes, whereas the later peak under the age of 60 years may reflect the higher proportions at these younger ages being admitted to critical care resulting in an extension of life of several days.

**Competing interests:** All authors have completed the ICMJE uniform disclosure form at www.icmje.org/coi_disclosure.pdf and declare: no support from any organization for the submitted work; no financial relationships with any organizations that might have an interest in the submitted work in the previous three years other than LS who reported grants from Wellcome, MRC, NIHR, GSK, BHF, Diabetes UK all outside the submitted work; no other relationships or activities that could appear to have influenced the submitted work other than LS who is a Trustee of the British Heart Foundation and AJM who is a member of the Royal Society Delve Committee.

## Background

Determining if and when the mortality peak of the current COVID-19 epidemic has been reached in any particular country is a key input into government, health service planning and guiding public health strategy. It is also crucial information to communicate to the public who in most countries have been subject to varying degrees of limitation and restriction on social interaction, work and movement (lockdown). While it is repeatedly stated by politicians and scientific experts that there is inevitably a delay between the point at which social distancing, individual protection and lock-down measures are imposed and the point when this may result in declining mortality, the uncertainty about when the daily toll of deaths is likely to decline will add to widespread public anxiety about the epidemic (1).

A full understanding of the dynamics of this and any other epidemic requires information on transmission, incidence of new cases and prevalence of active infection and immunity in the population (2). Reliable data of these epidemiological parameters is fragmentary in many countries. In the UK, the absence of large-scale testing for active infection has been regarded as a particular problem (3), with an almost exclusive focus until recently on testing among those admitted to hospital (4). Testing of people admitted to hospital with suspect COVID-19 infection is common in many countries and is recommended as a key priority within an overall testing strategy (2, 5). It provides a transparent and standardised metric of the epidemic burden : the numbers of people dying in hospital who were COVID-19 positive at the time of death. In the UK, the number of hospital deaths with COVID-19 is one of the headline figures provided daily, and the trajectory of the cumulative number of such deaths are the basis for many widely disseminated comparisons of how the UK is doing compared to others (6).

The interpretation of trends in the daily headline count of COVID-19 deaths in the UK is problematic for a number of reasons. One of the key challenges is that this number is comprised of COVID-19 deaths in hospital reported for the first time to the relevant authorities in a defined 24 hour period regardless of when each death actually occurred. The reasons for delayed reporting are several and will reflect pressures across the hospital system during the epidemic as well as pre-existing variation in quality and efficiency of data infrastructure. Thus for any particular reporting day, deaths are included whose reporting was delayed by 1, 2, 3 or more days. While this is clearly stated by NHS England on their website (7), this distinction appears to be lacking in most public debate and discussion relating to the figures.

In this paper we utilised datasets on COVID-19 deaths in hospital from NHS England that have been published by NHS England and announced to the public on a daily basis since early April to i) quantify the bias inherent in using the total number of such reported deaths as a metric of intensity and trajectory of mortality; ii) examine whether there is evidence of a downturn in hospital mortality once delays in reporting of these deaths are taken into account; iii) critically examine the strengths and weaknesses of these data as indicators of the burden of COVID-19 in the UK.

### Methods

Since April 4 on a daily basis the number of COVID-19 deaths occurring in hospital in England up until 5pm the previous day, are published by the NHS. COVID-19 deaths in this context are those that occur in hospital of patients that had tested positive for the virus at the time of death. They are reported centrally through the COVID-19 Patient Notification System. The headline figure that attracts attention in the media is the total number of deaths that are announced for the 24 hours up to 5pm the previous day. For example, on 14 April a total of 744 deaths were announced. These represent deaths reported to the centre in the period from 5pm 12 April to 5pm 13 April.

We downloaded the daily Excel spreadsheets from the Department of Health website (7) published each day from April 2 to April 25. These showed numbers of deaths reported by actual date of death for all ages combined, and separately for 5 age groups (0-19, 20-39, 40-59, 60-79 and 80+ years). Further breakdown by sex was not available. In our analyses we focus on all ages and the three older age groups as the number of deaths under the age of 40 was very small and thus not meaningful to analyse separately. For the whole of the epidemic up until the data published on April 25 only 139 of the total of 18084 deaths were in this youngest age group.

In the first analyses we examine the delay in reporting of COVID-19 hospital deaths and look at how this can bias impressions about trends. We then present analyses that aim to provide a less biased impression of the trajectory of the epidemic in so far as it is measured by daily counts of COVID-19 deaths in hospital. We did this by looking at the trend over time in the numbers of deaths reported within a particular period of delay. While the absolute numbers of deaths seen with 1, 2 or even 5 days delay represent an underestimate of the total deaths occurring on any one day, focussing on trends within single days of delay periods allows a fairer, more like with like comparison of death counts by date of death.

## Results

Figure 1 shows how the COVID-19 deaths published by NHS England on one of four arbitrarily selected days are comprise of deaths that actually occurred over a span of a wide range of previous days. For any one day of death, the maximum number of deaths are announced with a delay of 2 days. An appreciable number of deaths are included with greater delays. Over the reporting period from April 2 to April 20, on average 90% of deaths announced were included after 5-days. What is also evident from Figure 1 is that over time that has been a slight but noticeable decline in the delays of reporting, with a tighter and more symmetrical distribution of deaths by delay evident for daily announced deaths up to April 20.

**Figure 1:**
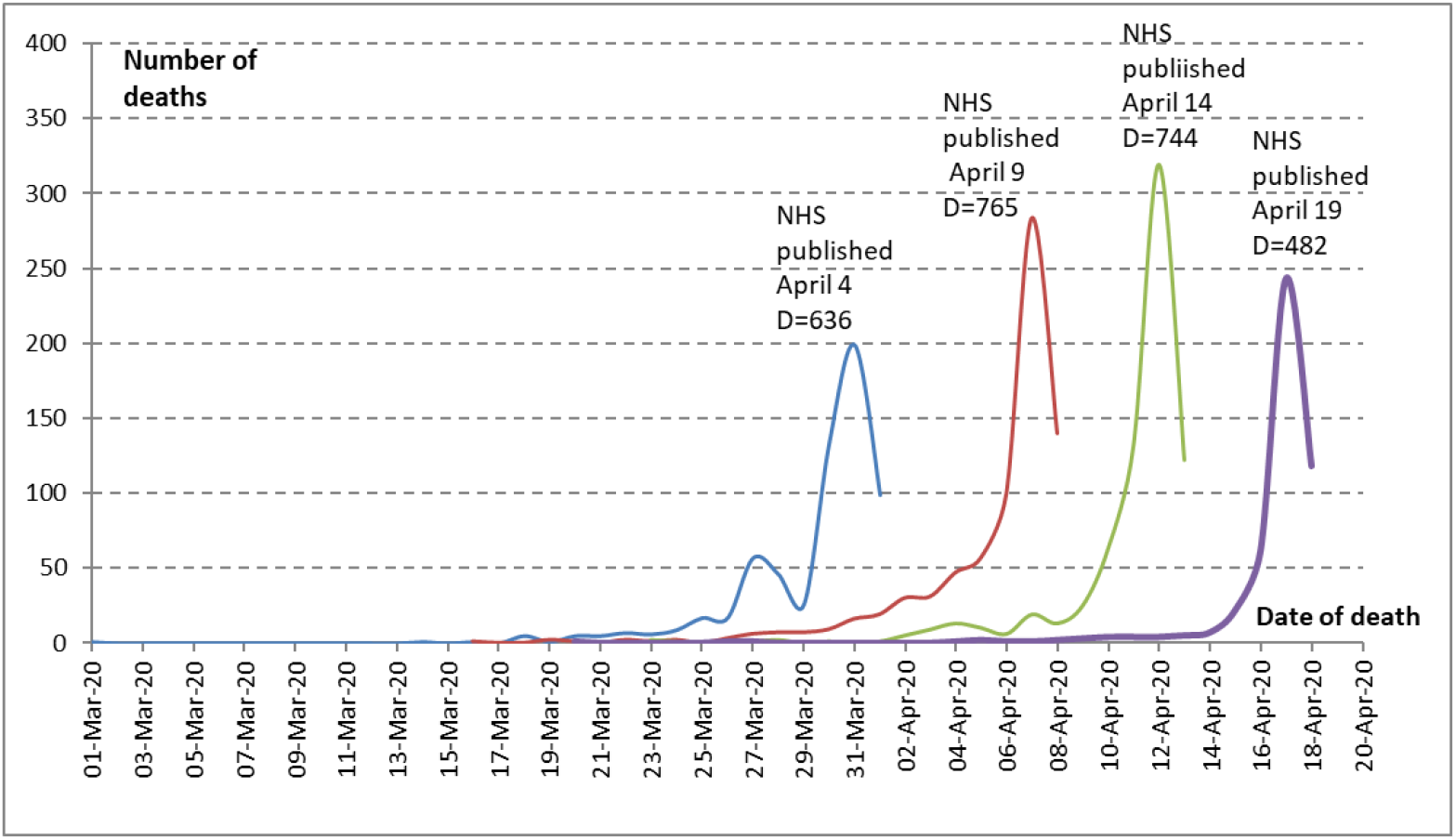
Distribution of hospital COVID-19 deaths by date of death announced by NHS England on selected dates.

Figure 2 addresses the extent to which the total number of daily deaths published by the NHS provides a somewhat misleading impression of levels of and trends in actual daily mortality based on the date of death per se. Looking at the maximum level of daily announced deaths (blue line) this has a maximum (866 on April 10) that is higher than the observed maximum of daily deaths (red line) by date of occurrence (801 on April 8). More importantly, the trajectory of the daily announced deaths is more unstable than the trajectory shown for deaths by date of occurrence. Specifically, there is an indication that the maximum total number of hospital deaths occurred on April 8, which is just not evident when looking at trends in daily announced deaths.

**Figure 2:**
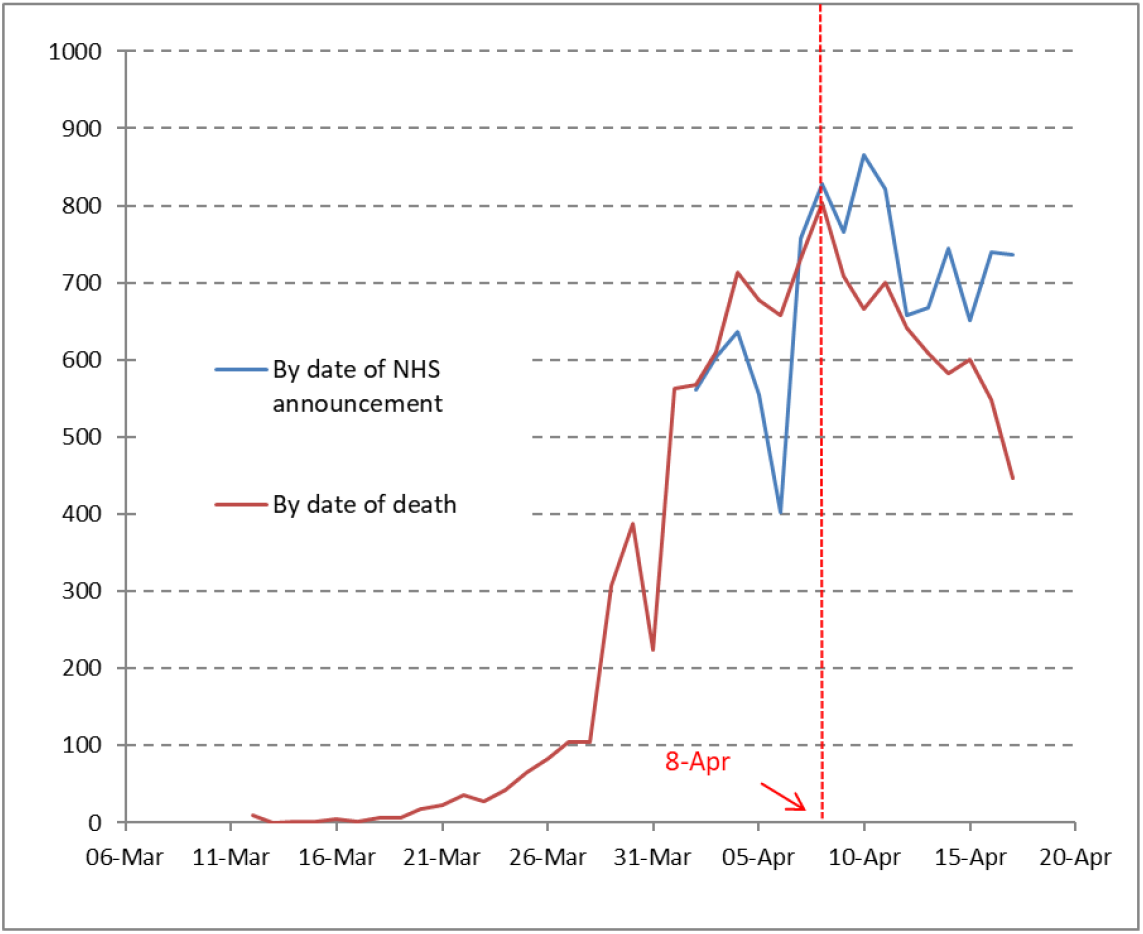
Numbers of published hospital COVID-19 daily deaths* according to date of announcement by NHS England and actual date of death (up to April 17) *Note : the deaths included in this Figure are only those included in the daily reports announced between April 2 and April 20. The report for April 2 only included those deaths occurring at an earlier date that had been notified to NHS England from 5pm 31 March to 5Pm 1 April

Figure 3 provides further insights into how the correspondence between the dates of the announcement and the dates of occurrence affects the temporal pattern of deaths at all ages and in specific age groups. It shows for all deaths and those in selected age groups trends in total numbers of deaths that occurred on any particular day that were reported with delays of between 1 and 7 days. The numbers on which these Figures are based are shown in Table 1. Note that the numbers of deaths shown as occurring for any one day for a given delay are the sum of all deaths on that day cumulated across preceding delay periods. For example, looking at all deaths in panel (a) one can see that there were 140 deaths on April 8 that were reported with 1 day’s delay (i.e. announced on April 9). However, taking account of deaths reported with both 1 and 2 day’s delay, the number of deaths seen for April 8 rose to 496.

**Table 1:**
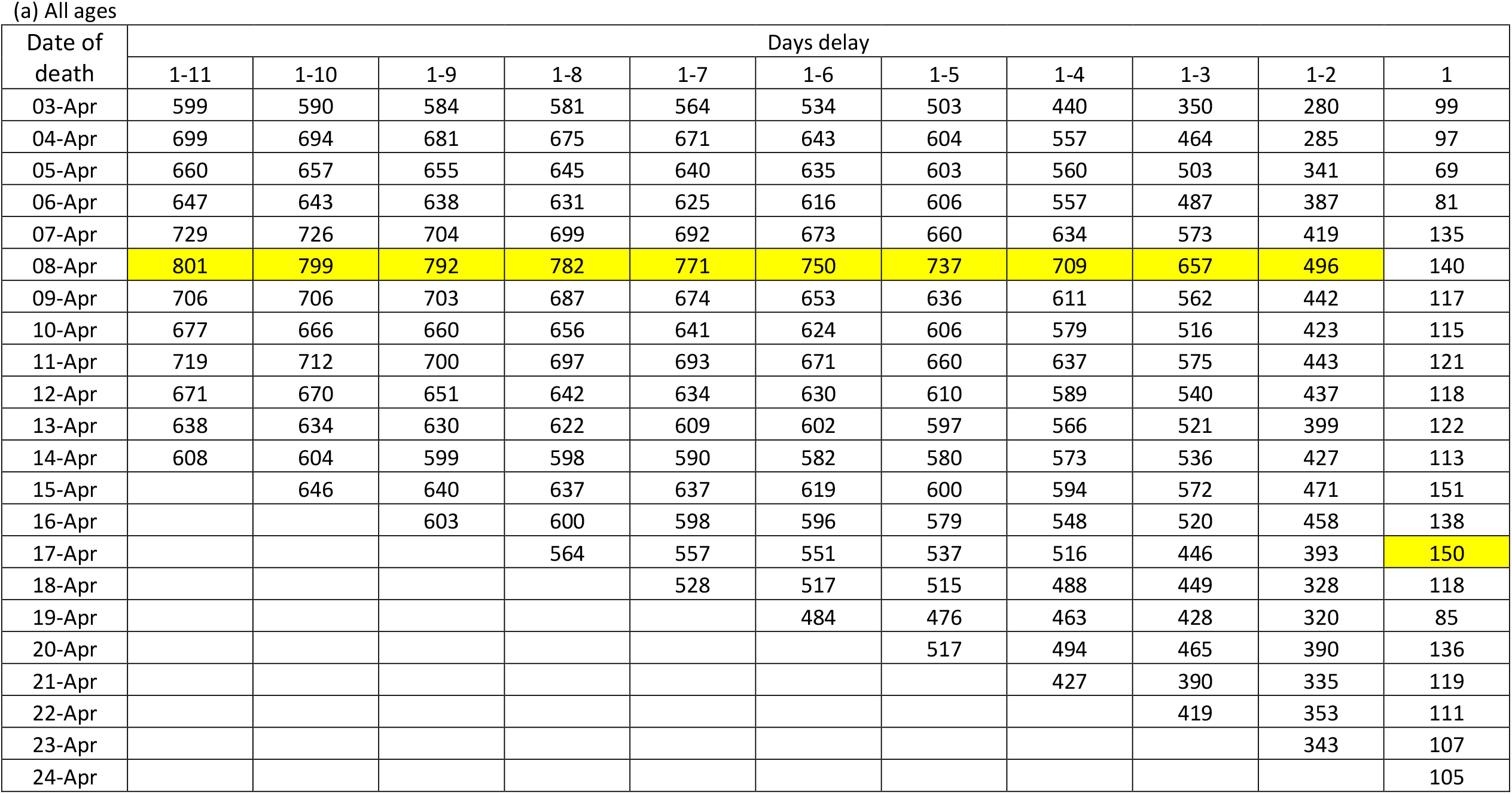
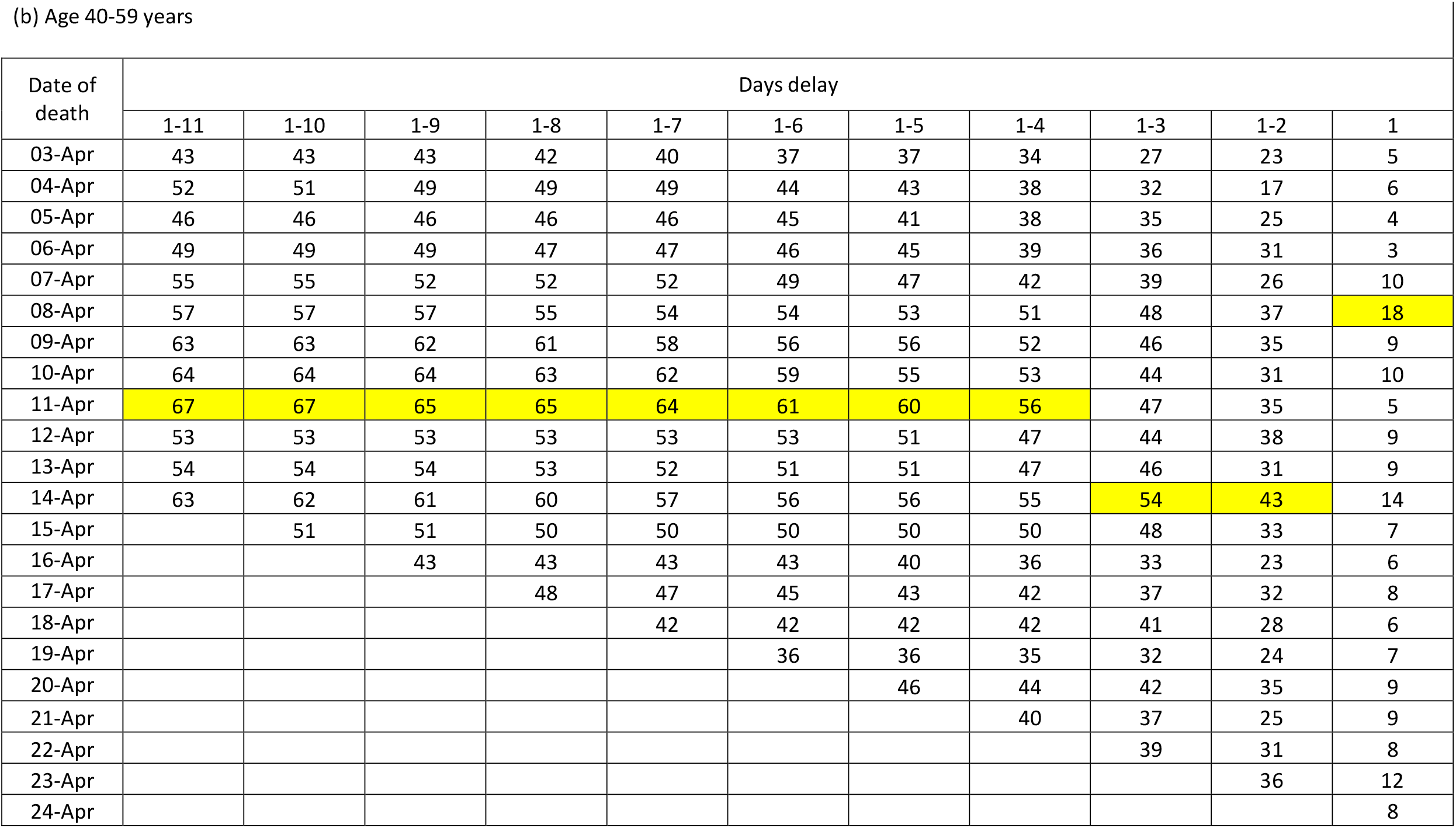
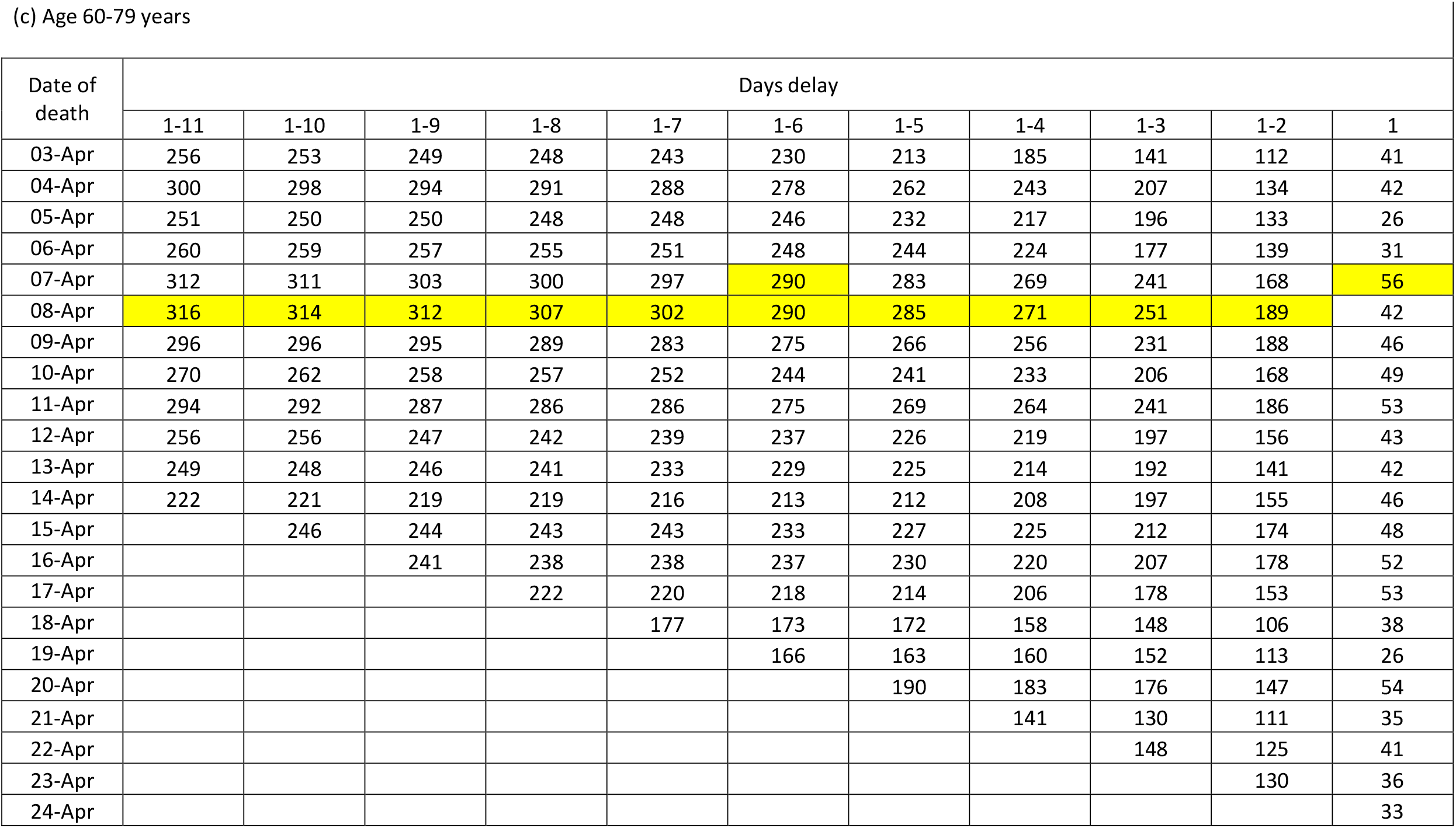
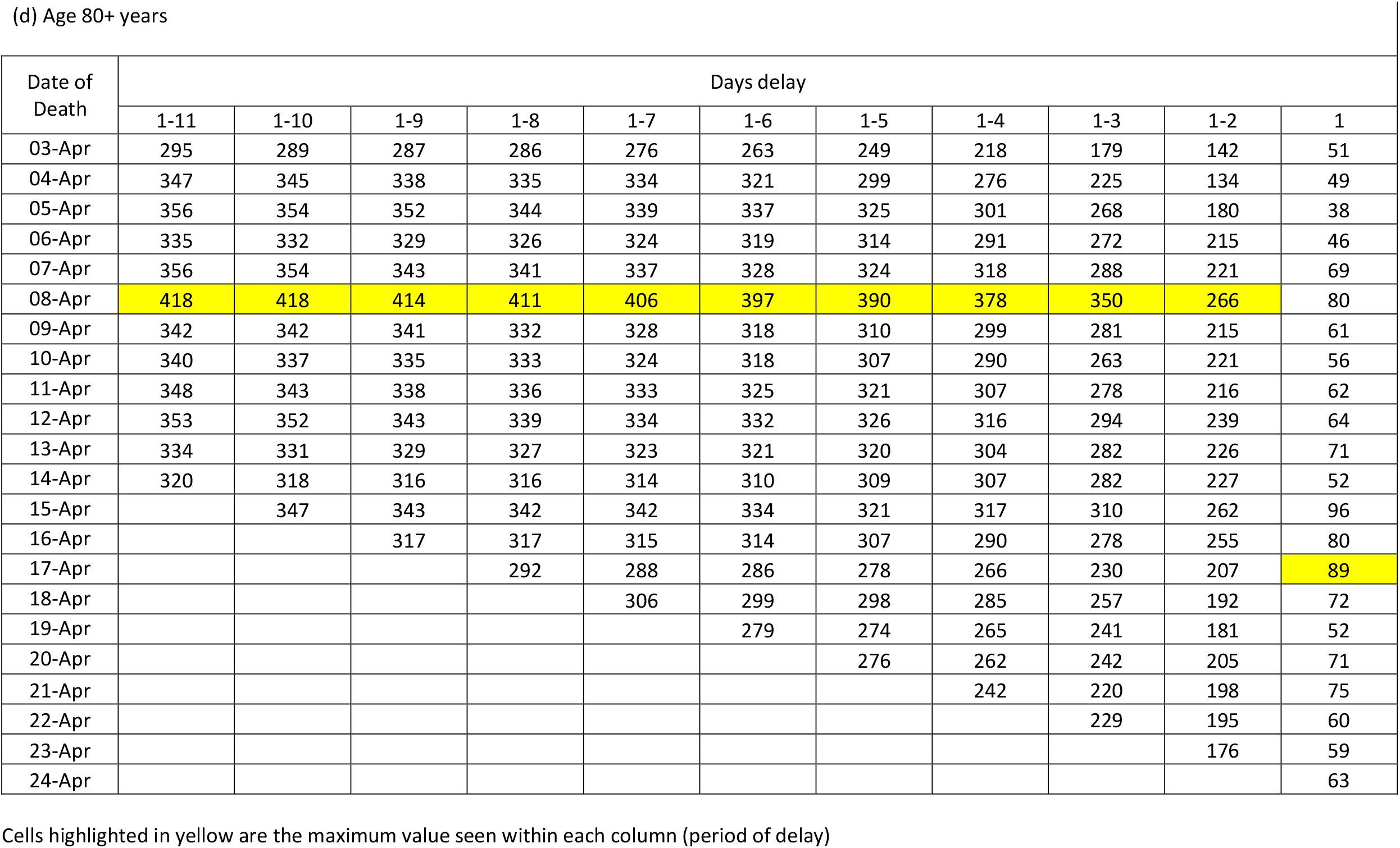
Cumulated numbers of hospital COVID-19 deaths by date of death (up to April 24) according to length of delay (in days) between this date and date of announcement by NHS England (up to April 25), by age at death

**Figure 3:**
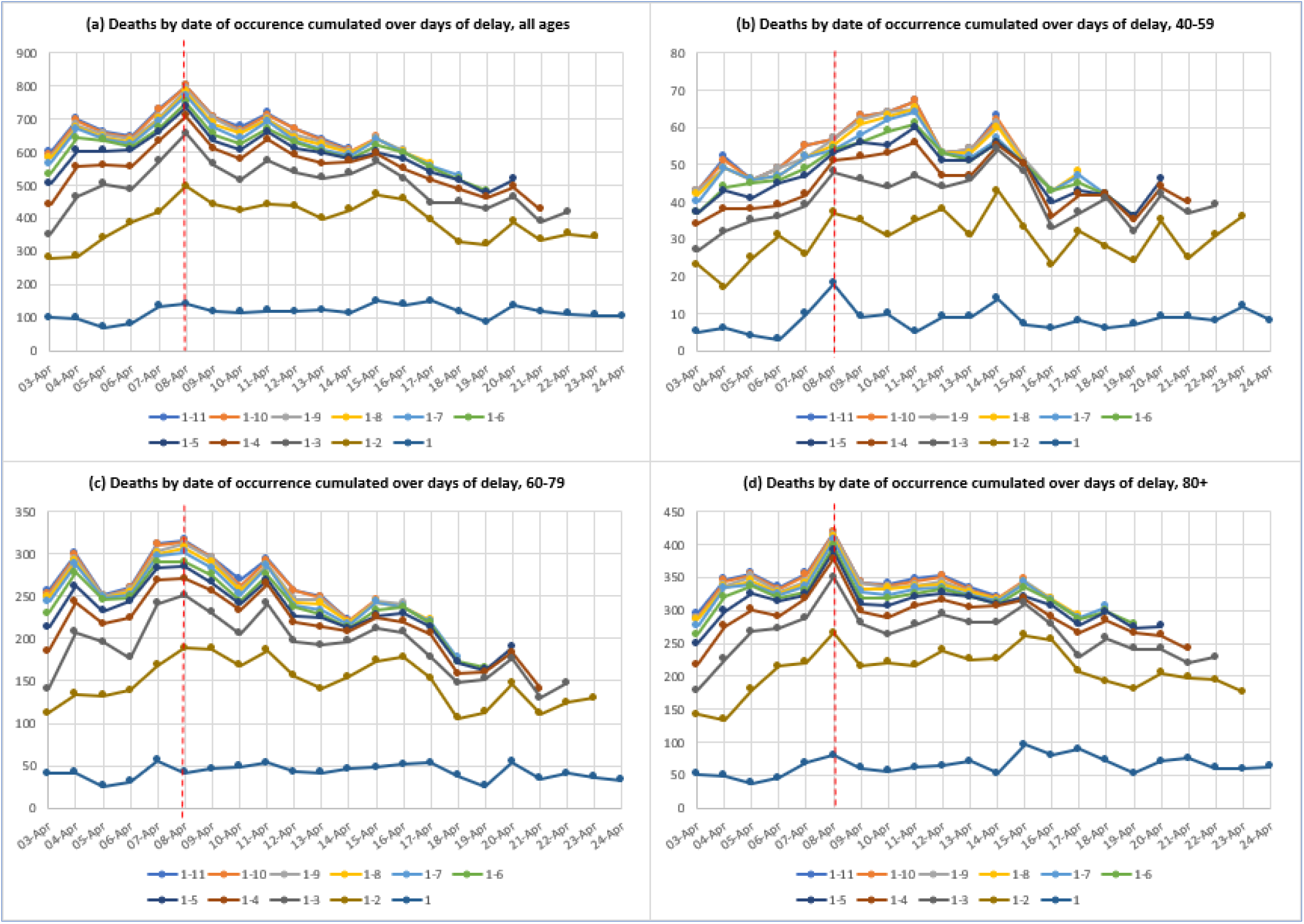
Numbers of hospital COVID-19 deaths by date of death (up to April 24) according to length of delay (in days) between this date and date of announcement by NHS England (up to April 25). Shown for all ages combined and selected adult age groups separately. Note : vertical dashed red line indicates deaths occurring on 8 April 2020

Examination of the overall pattern seen for all deaths (Figure 3, panel (a)) reveals that the highest number of deaths within each delay period is April 8 other than for 1 day’s delay. This is confirmed in Table 1. From April 8 there is good evidence of a decline, with a small perturbation on April 11. For deaths aged 40-59 years (panel (b)) the peak appears to occur 3 days later on April 11. However, at ages 60-79 and 80+ the April 8 peak is evident, although as in the youngest age group there is evidence of a small peturbation on April 11. The oldest age group is notable in that although the peak on April 8 is clear, there is less evidence of a decline after April 9, with indications of a plateau having been reached.

The patterns by English region vary. Equivalent plots and numbers are given in Figure 4 and Table 2 for London and the Midlands. The picture for London is of a convincing decline since April 8, although there is evidence of a slightly higher peak earlier than this on April 4. In contrast, while in the Midlands there is a peak on April 8, the subsequent decline is not as smooth as in London, with a further smaller peak on April 15.

**Table 2:** Cumulated numbers of hospital COVID-19 deaths by date of death (up to April 24) according to length of delay (in days) between this date and date of announcement by NHS England (up to April 25) for London and the Midlands

| Date of death | Days delay |  |  |  |  |  |  |  |  |  |  |
| --- | --- | --- | --- | --- | --- | --- | --- | --- | --- | --- | --- |
|  | 1-11 | 1-10 | 1-9 | 1-8 | 1-7 | 1-6 | 1-5 | 1-4 | 1-3 | 1-2 | 1 |
| 03-Apr | 165 | 159 | 156 | 155 | 149 | 139 | 124 | 112 | 84 | 62 | 15 |
| 04-Apr | 200 | 198 | 191 | 189 | 189 | 178 | 165 | 149 | 127 | 79 | 19 |
| 05-Apr | 163 | 161 | 161 | 157 | 154 | 154 | 140 | 128 | 116 | 84 | 12 |
| 06-Apr | 164 | 164 | 161 | 159 | 159 | 155 | 150 | 137 | 123 | 98 | 15 |
| 07-Apr | 189 | 187 | 176 | 173 | 171 | 167 | 163 | 153 | 140 | 90 | 29 |
| 08-Apr | 199 | 199 | 197 | 193 | 190 | 182 | 180 | 170 | 156 | 125 | 29 |
| 09-Apr | 173 | 173 | 172 | 160 | 159 | 153 | 151 | 144 | 134 | 110 | 31 |
| 10-Apr | 149 | 149 | 149 | 146 | 144 | 141 | 134 | 126 | 101 | 87 | 29 |
| 11-Apr | 158 | 156 | 156 | 156 | 156 | 148 | 147 | 140 | 124 | 87 | 13 |
| 12-Apr | 148 | 147 | 132 | 132 | 132 | 132 | 130 | 127 | 114 | 100 | 22 |
| 13-Apr | 148 | 148 | 147 | 142 | 142 | 142 | 141 | 128 | 120 | 88 | 24 |
| 14-Apr | 122 | 121 | 121 | 120 | 117 | 116 | 115 | 114 | 103 | 78 | 28 |
| 15-Apr |  | 131 | 130 | 130 | 130 | 117 | 114 | 112 | 108 | 76 | 20 |
| 16-Apr |  |  | 130 | 130 | 130 | 129 | 121 | 119 | 108 | 92 | 25 |
| 17-Apr |  |  |  | 88 | 88 | 87 | 81 | 80 | 73 | 61 | 27 |
| 18-Apr |  |  |  |  | 93 | 91 | 90 | 82 | 77 | 56 | 21 |
| 19-Apr |  |  |  |  |  | 95 | 94 | 92 | 87 | 67 | 12 |
| 20-Apr |  |  |  |  |  |  | 79 | 77 | 71 | 60 | 20 |
| 21-Apr |  |  |  |  |  |  |  | 80 | 72 | 51 | 26 |
| 22-Apr |  |  |  |  |  |  |  |  | 90 | 72 | 16 |
| 23-Apr |  |  |  |  |  |  |  |  |  | 59 | 13 |
| 24-Apr |  |  |  |  |  |  |  |  |  |  | 9 |

Cells highlighted in yellow are the maximum value seen within each column (period of delay) (b) Midlands
| Date of death | Days delay |  |  |  |  |  |  |  |  |  |  |
| --- | --- | --- | --- | --- | --- | --- | --- | --- | --- | --- | --- |
|  | 1-11 | 1-10 | 1-9 | 1-8 | 1-7 | 1-6 | 1-5 | 1-4 | 1-3 | 1-2 | 1 |
| 03-Apr | 117 | 117 | 117 | 116 | 114 | 101 | 96 | 85 | 68 | 39 | 15 |
| 04-Apr | 135 | 135 | 135 | 131 | 128 | 127 | 114 | 107 | 88 | 35 | 3 |
| 05-Apr | 148 | 147 | 146 | 146 | 145 | 144 | 137 | 120 | 107 | 49 | 0 |
| 06-Apr | 132 | 132 | 132 | 132 | 132 | 128 | 125 | 111 | 87 | 65 | 7 |
| 07-Apr | 116 | 116 | 116 | 116 | 116 | 116 | 112 | 111 | 103 | 55 | 14 |
| 08-Apr | 173 | 173 | 173 | 173 | 173 | 171 | 169 | 163 | 148 | 92 | 17 |
| 09-Apr | 130 | 130 | 130 | 128 | 127 | 124 | 119 | 117 | 102 | 72 | 12 |
| 10-Apr | 124 | 124 | 124 | 124 | 118 | 115 | 115 | 110 | 106 | 71 | 14 |
| 11-Apr | 137 | 136 | 136 | 136 | 136 | 132 | 130 | 127 | 113 | 92 | 13 |
| 12-Apr | 132 | 132 | 132 | 131 | 129 | 128 | 124 | 119 | 110 | 87 | 19 |
| 13-Apr | 113 | 113 | 111 | 110 | 110 | 110 | 109 | 98 | 88 | 70 | 12 |
| 14-Apr | 114 | 114 | 114 | 114 | 113 | 113 | 113 | 112 | 106 | 80 | 19 |
| 15-Apr |  | 144 | 144 | 144 | 144 | 143 | 143 | 143 | 141 | 116 | 30 |
| 16-Apr |  |  | 92 | 91 | 89 | 89 | 85 | 83 | 81 | 73 | 8 |
| 17-Apr |  |  |  | 112 | 112 | 112 | 108 | 103 | 94 | 79 | 22 |
| 18-Apr |  |  |  |  | 110 | 110 | 109 | 107 | 95 | 61 | 20 |
| 19-Apr |  |  |  |  |  | 88 | 88 | 87 | 79 | 60 | 6 |
| 20-Apr |  |  |  |  |  |  | 103 | 99 | 95 | 72 | 21 |
| 21-Apr |  |  |  |  |  |  |  | 78 | 75 | 64 | 16 |
| 22-Apr |  |  |  |  |  |  |  |  | 66 | 54 | 14 |
| 23-Apr |  |  |  |  |  |  |  |  |  | 78 | 15 |
| 24-Apr |  |  |  |  |  |  |  |  |  |  | 15 |

Cells highlighted in yellow are the maximum value seen within each column (period of delay)

**Figure 4:**
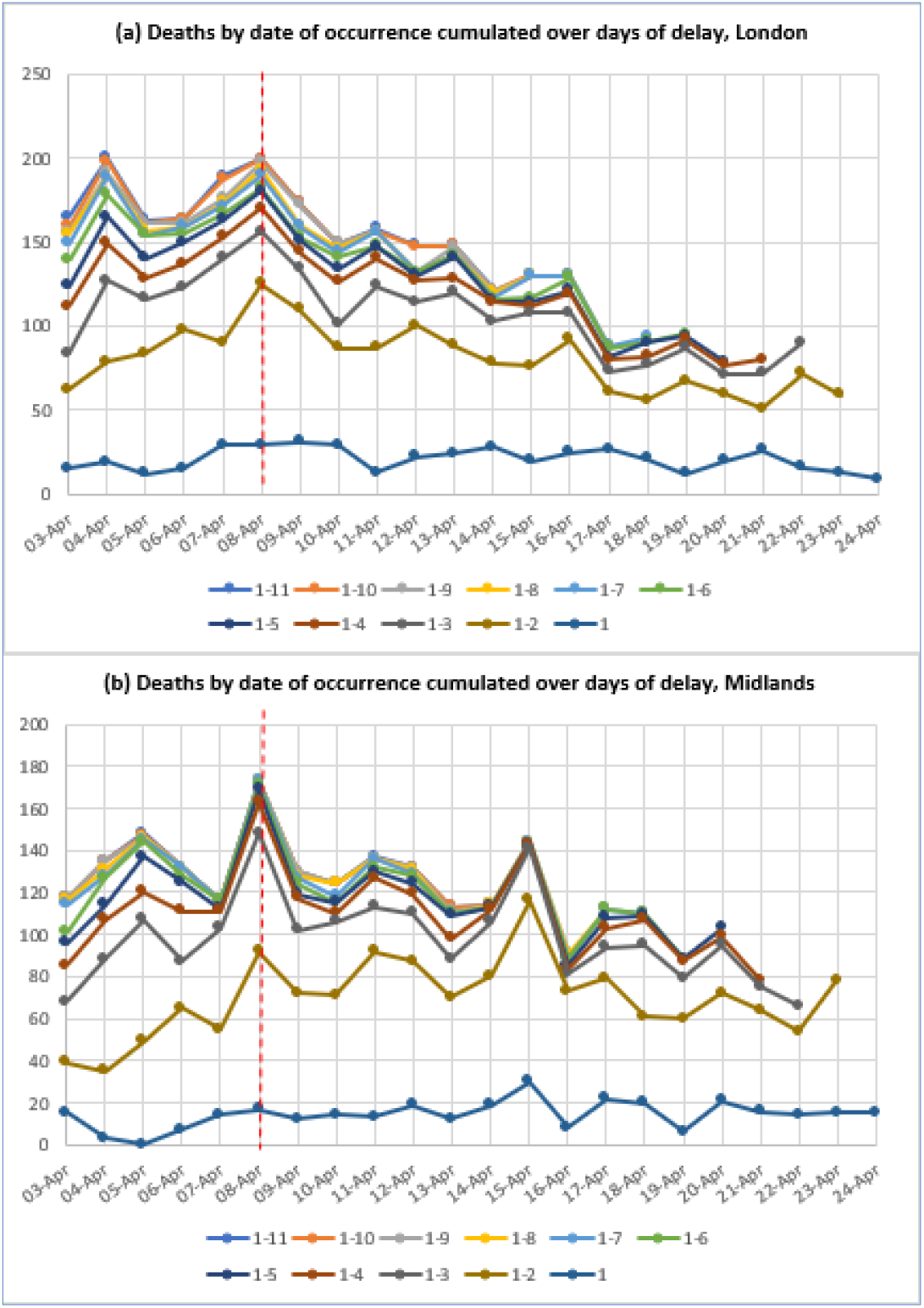
Numbers of hospital COVID-19 deaths by date of death (up to April 24) according to length of delay (in days) between this date and date of announcement by NHS England (up to April 25). Shown for London and the Midlands separately. Note : vertical dashed red line indicates deaths occurring on 8 April 2020

## Discussion

In this paper, we have illustrated the problems and biases inherent in using the total number of daily COVID-19 deaths published by NHS England and announced to the public as a guide both to the trajectory as well as the level of daily deaths by date of occurrence. Our analysis that makes a more like with like comparison over days by taking account of delay periods suggests that there may have been a peak of hospital COVID-19 deaths on April 8. This is consistent with the results of a more formal statistical approach to dealing with the problem of delay (8). However, this pattern varies somewhat by age and by region. Whether this really is the case will become clear when several more days of data have become available.

As we discuss below there are a number of caveats concerning how far hospital COVID-19 deaths can be regarded as reflecting the trajectory of the epidemic above and beyond the delays in reporting to the NHS that we have already discussed. However, as a first approximation, one may work back from a posible peak on April 8 to when the rate of community transmission began to fall. If we assume a median time of around 23 days following infection (5 days median incubation period (9) and 18 days from symptom onset to death(10)) one can track back to a date of infection of around March 16. This is a week prior to the hard lockdown announced on 23 March. However there is evidence of a decline in social contact and travel in the previous week (March 16-23) (11, 12).

From another perspective, this illustrates just how long the delay is between a fall in rate of infection and it becoming manifest in a fall in deaths. To the 23 days one would need to add an additional 5 or more to be confident that the mortality signal was not due to delays in reporting, resulting in a 28 day gap between a decline in infections becoming apparent in deaths by date of occurrence.

The dynamics of infection and subsequent mortality are importantly driven by changes in patterns of transmission in the community. However, it is important to note that community transmission is not the only source of infections that lead to death in hospital. There are also infections within care-homes, which will behave differently and will not be as influenced by broader lockdown once established in any particular institution. Although care homes will have introduced restrictions on visitors who may seed infection, they are not entirely closed communities, and once established in a particular institution, transmission may be particularly difficult to stop. Lockdown primarily affects inter-household and not intrahousehold transmission. This may lead to a more extended period of ongoing transmission and later mortality than seen in the general community. Finally there are the infections that occur within hospitals, that will affect both staff as well as others who have to be in hospital for other reasons other than COVID-19. Again, this focus of transmission will have its own dynamic.

The most striking finding by age concerns those dying aged 40-59 years, who look as though they have a peak mortality on April 11 rather than April 8. Because they constitute less than 1 in 10 of total hospital COVID-19 deaths, the effect of this later peak is not very visible in the pattern seen for the population as a whole. One possible explanation for this later peak is that a larger proportion of these younger cases who die have been admitted to critical units than at older ages. Although they eventually succumb to the infection, their survival might have been extended by a few days as a result of the more invasive ventilation and organ support available in critical care units. This supposition is supported by a comparison of the age distributions of patients with COVID-19 admitted to critical units who subsequently die, compared to the age distribution of all hospital COVID-19 deaths. Data from a recent report of the Intensive Care National Audit & Research Centre on COVID-19 patients (13) shows that up to April 16 only 2% of the COVID-19 deaths in critical care units were to people aged 80+. In comparison 52% of all hospital COVID-19 deaths are among those aged 80+ up to the same date. Indeed only 3% of the COVID-19 cases admitted to critical care were aged over 80 years, while 46% were younger than 60 years.

The reasons why London shows a clearer pattern of decline after April 8 compared to the country as a whole or the Midlands are unclear. It is well accepted that the epidemic in London was more advanced and intense than in other regions. It is possible that the effect of the lockdown on transmission via the London underground and other features of the human density of the city may have been sharper than in other places. However, further work is required to understand this.

Our analyses are based on counts of people who die in hospital with a laboratory verified diagnosis of SARS-CoV-2. This definition of a COVID-19 death is similar to that used in a number of other countries including Italy (14). However not all of these deaths will be caused by COVID-19, although it seems likely a large number will have been precipitated by the infection. However, more importantly, there is the issue of deaths occurring outside of hospital. The weekly ONS COVID mortality report (published April 21) showed that to April 10 18% of deaths in which COVID-19 was mentioned on the death certificate occurred in private homes, care homes or hospices (15). It is, however, unclear what proportion of cases of people in care homes are admitted to hospital if they have COVID-19 symptoms. Crucially for the validity of our analyses it is not known whether the fraction of such cases admitted has changed during the course of the epidemic. On the one hand as the epidemic has progressed there may be a greater reluctance to send to hospital frail elderly people with presumed COVID-19. On the other hand the probable steep growth in the number of cases occurring in care homes in April, as evidenced by a doubling of deaths mentioning COVID-19 on the death certificate over the week 10 April (15), suggests that there might have been an increase in admissions from this source. These two forces may balance out as suggested by the fact that there is a shallower decline in hospital COVID-19 deaths at age 80+ years after the April 8 peak compared to the age group 60-79 years.

A final caveat is important. Not all of the hospital COVID-19 deaths we have analysed will have been caused by COVID-19. A subset will really be people admitted to hospital and critical care units with other life threatening conditions including terminal cancer and trauma (16). It is quite conceivable that a fraction of these will be infected with COVID-19 in the hospital itself. These deaths will however be still classed as COVID-19 using the pragmatic definition used to assemble these data. Many of these deaths are however unlikely to be coded as having COVID-19 as the underlying cause of death.

What are the implications of our findings? For analytic purposes, the sorts of analyses we have undertaken (Figure 3) where counts are examined within defined single-day periods of delay provide a more sensitive and transparent approach to the empirical analysis of the trends. This will be particularly important to consider when making international comparisons, where other patterns of delay may have occurred. With respect to public communication of trends we suggest that emphasis is placed upon trends in deaths by date of occurrence that are evident with 5 or more days delay.

Establishing when the peak of COVID-19 mortality occurred will help anchor modelling of the so far hidden extent of the epidemic infection in England, and will also help with health service planning. The fact that a peak may have been reached on April 8 is not an argument for saying that restrictions on social contact and so on should now be relaxed. Communicating to the public that there has been one peak a week ago might provide in fact motivation for people to persevere with the lockdown strategy – as even deaths are responding to its imposition.

## Data Availability

All data analysed are in the public domain. URLs are provided for all data sources.

## Notes

### Competing Interest Statement

LS reported grants from Wellcome, MRC, NIHR, GSK, BHF, Diabetes UK all outside the submitted work; LS reported being a Trustee of the British Heart Foundation and AJM reported being a member of the Royal Society Delve Committee.

### Funding Statement

This work was done without specific funding

